# Identifying Non-traditional Epidemic Disease Risk Factors associated to major Health Events from WHO and World Bank Open Data

**DOI:** 10.1101/2020.06.10.20127746

**Authors:** Roberta Lugo-Robles, Eric C. Garges, Cara H. Olsen, David M. Brett-Major

**Affiliations:** Uniformed Services University, Bethesda, Maryland, USA; Henry M. Jackson Foundation, Bethesda, Maryland, USA; University of Nebraska Medical Center, Omaha, Nebraska, USA

## Abstract

**Objectives:** Health events emerge from a multifactorial milieu involving host, community, environment, and pathogen factors. Therefore, developing accurate forecasting models to improve epidemic prediction towards better prevention and capabilities management is a complex task. Here, we describe an exploratory analysis to identify non-health risk factors that could improve the forecast and events risk signals using a feasible and practical approach by combining surveillance report data with non-health data from open data sources.

**Methods:** A line listing was developed using information from the World Health Organization Disease Outbreaks News from 2016-2018. A database was created merging the line listing data with non-health indicators from the World Bank. Poisson regression models employing forward imputations were used to establish relationships and predict values over the dependent variable (health event frequency); which are the health events reported by each country to WHO during 2016-2018.

**Findings:** The resulting regression model provided evidence that changes in non-health factors important to community experiences impact the risk of the number of major health events that a country could experience. Three non-health indicators (extrinsic factors) were associated significantly to event frequency (population urban change, gross domestic product change per capita—a novel factor, and average forest area). An exploratory analysis of the current COVID-19 pandemic suggested similar associations, but confounding by global disease burden is likely.

**Conclusion:** Continued development of forecasting approaches capturing available whole-of-society extrinsic factors (non-health factors); could improve the risk management process through earlier hazard identification, and as importantly inform strategic decision processes in multisectoral strategies to preventing, detecting, and responding to pandemic-threat events.

## Introduction

The challenge of employing infectious disease outbreak data usefully to help those managing an emergency, and the need to purposefully develop models oriented to decision makers and in a context of being connected to management, is increasingly recognized.(1) There are several barriers to meeting these. Among them are availability of validated data before an emergency to proof the model, and during an event to give actionable information. Such efforts focus on early warning of an event or its trajectory. They often are anchored on specific characteristics of how an outbreak pathogen behaves. This may result in clumsy applications of information oriented in that way to a community-based perspective of what must be done to prevent and mitigate risks. Here, we attack these challenges from the flank, pursuing in a pilot analysis prediction in a broad stroke across many pathogens rather than early warning; seeking to demonstrate potential utility of openly available information from both health and non-health inter-agencies; and, exploring triggers meriting prevention and early mitigation efforts rather than response.

The Disease Outbreak News (DON) by the World Health Organization is a major conduit for information sharing relevant to state party obligations under the International Health Regulations. Last published in 2007, the International Health Regulations includes an assessment and notification tool to report events that may constitute a public health emergency of international concern.(2) The outputs from this risk identification tool are translated into brief reports for the DON. Each WHO Member State is meant to report events in accordance to these Regulations.(2)

We aimed to 1) describe reported outbreak events impactful to communities via the WHO Disease Outbreak News from 2016-2018; 2) identify and explore major extrinsic factors at the population level potentially related to these events; and, 3) generate hypotheses regarding associations between extrinsic factors and events for the purpose of identifying triggers for enhanced surveillance, other health system strengthening, or holistic community interventions that might later be investigated for either early risk mitigation or prevention of pandemic-threat events. Detecting early risk signals boosts the risk management process (Figure 1) through longer lead time for risk identification and characterization.

**Figure 1.**
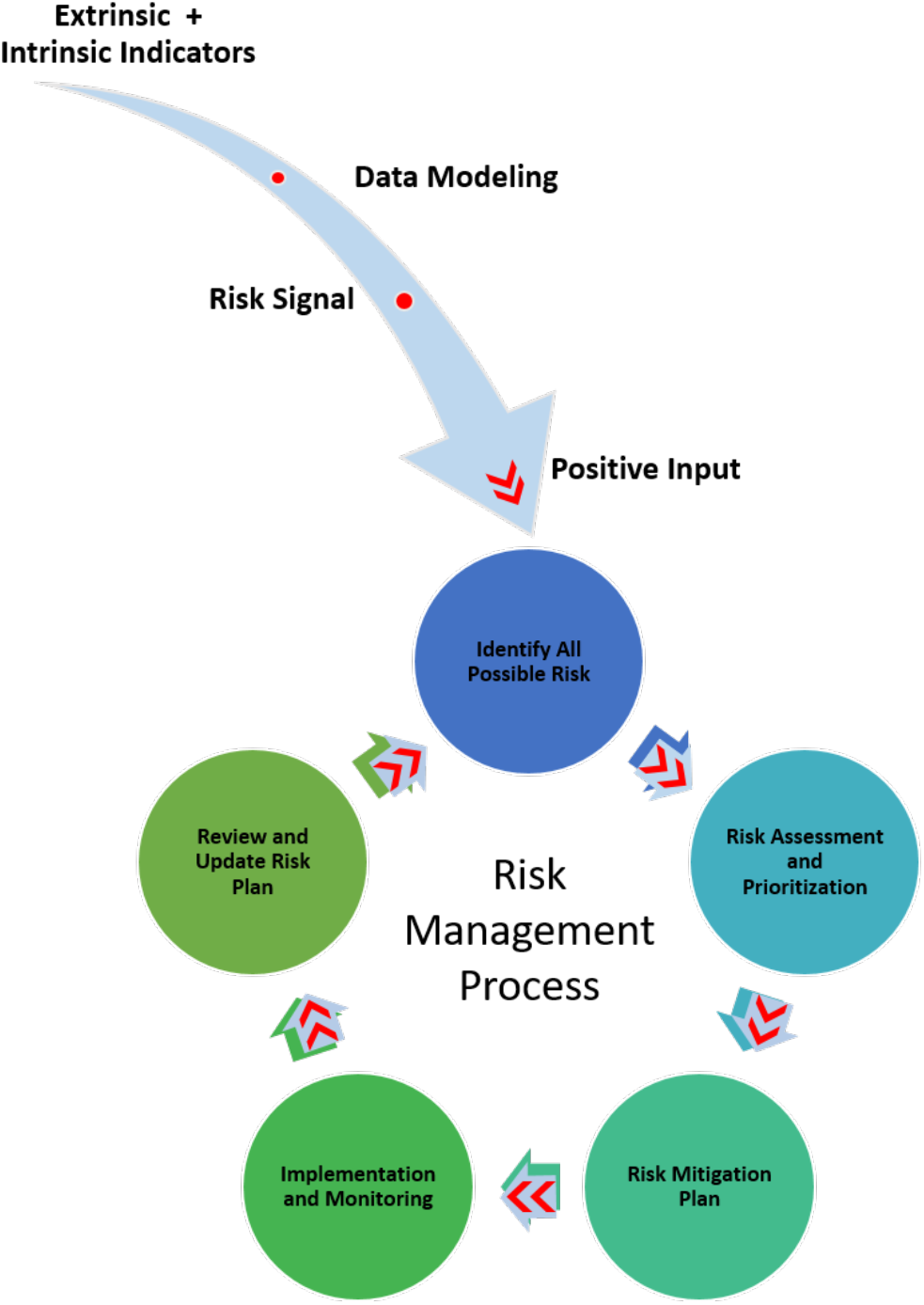
Risk signal identification and input effect on the risk management process.

## Methods

Information contained in WHO Disease Outbreaks News from 2016-2018 was reviewed, and employed to construct a line listing. Each row of the line listing identified a single outbreak (event) in a single country. Multiple reports corresponding to the same outbreak (or event) were summarized within the same row. Outbreak information such as demographics and epidemiological indicators were recorded, as available.

Extrinsic factors were defined as non-biological factors that could contribute to the risk of a health event regardless of the pathogen involved. The main reason to understand extrinsic factors and health events is to account for predictability. However, as outbreaks spread and spatiotemporally separated waves become entangled with human mobility, behavioral changes, pathogen evolution and other factors, the predictability of the models structures decrease despite increasing time-series lengths.(3) This make the prediction of an event very complex. We limited our study to evaluate a specific amount of predictors to explore extrinsic factors associated with frequency of health events. In particular, we employed World Bank’s World Development Indicators. They are internationally comparable statistics about global development selected as community-centric risk factors which are routinely assessed and could inform a risk management and prioritization process. We selected particular factors for their potential relevance to communities’ vulnerability and exposure. Together with health event data, this information combined in a model will served as a proxy to predict health events frequency (risk signals) in a feasible and practical approach aim to impact the risk management process.

### Data Management

Data was aggregated using country (N=96) as a grouping variable. Events’ frequency (dependent variable) was summed and grouped per country. All the countries reported at least one event within the timeframe analyzed. Independent variables related to economics, population level and environment were collected at baseline and current years. Baseline values were considered 10 years before (2006-2008) the DON reports included in the line list (2016-2018). Independent indicators were extracted from the World Bank open data repository. This database contains 1,600 time series indicators for 217 economies and more than 40 country groups from the last 50 years.(4)

### Statistical Analysis

For each independent variable an average delta (Δ) was calculated as the difference between averages per annum baseline values (2006-2008) and average current per annum values (2016-2018). Descriptive statistics were used to describe the health events in terms in terms of age, sex, case count, death count and other variables. Nonparametric statistics were applied to the dataset. Bi-variate analyses exploring associations included Spearman’s Rank Correlation Coefficient and Mann–Whitney U tests. Country comparisons to identify a potential confounding factor related to economic reporting behaviors relevant to achieving aid was performed. Data related to global health security funding commitments from 2014-2019 for the three countries in our data set with the most and least absolute gross domestic product (GDP) change was retrieved and compared using the Georgetown Infectious Disease Atlas.(5)

Poisson regression models employing forward imputations were used to establish relationships and predict values over the dependent variable. Model fitting was assessed by Omnibus Test (p<0.001) and the goodness of fit. Over-dispersion was evaluated by Pearson chi-Square value from the goodness of fit. All statistics were considered significant at p-value of 0.05. A Monte Carlo simulation was used using the predicting model equation obtained from the Poisson regression model to explore model performance.All input variables were fitted before running the simulation model.

## Results

From 2016 -2018, a total of 96 countries reported to WHO through the Disease Outbreak News the amount of 155 health events from 29 different pathogens (Figure 2a). The top five pathogens/ disease events reported are displayed in Figure 2b. The Zika outbreaks represented the 18.7% of the health evens reported, followed by MERS-CoV (11.0%) and Yellow Fever (8.4%). A Dengue virus event was the largest reported with more than 94,000 cases, followed by cholera with 27,978 cases. The demographic characteristics of the countries with more health events and least health events reported are depicted on Table 1. We observed no overall trend when comparing demographic indicators between countries with the highest number of reported health events against those with the number reported (a single report).

**Table 1.** Countries Demographic Characteristics for the year 2018

| Country | Health Events | Population | Mortality rate, under-5 (per 1,000 live births) | Birth rate, crude ( per 1,000 people) | Death rate, crude ( per 1,000 people) |
| --- | --- | --- | --- | --- | --- |
| Nigeria | 12 | 195,874,740 | 120 | 38 | 12 |
| Democratic Republic of Congo | 7 | 84,068,091 | 88 | 41 | 9 |
| China | 6 | 1,392,730,000 | 9 | 11 | 7 |
| United States | 6 | 326,687,501 | 7 | 12 | 9 |
| France | 5 | 66,977,107 | 4 | 11 | 9 |
| Cuba | 1 | 11,338,138 | 4 | 10 | 9 |
| Dominica | 1 | 71,625 | 36 | 12 | 8 |
| Guyana | 1 | 779,004 | 30 | 20 | 7 |
| United Kingdom | 1 | 66,460,344 | 4 | 11 | 9 |
| Malaysia | 1 | 31,528,585 | 8 | 17 | 5 |

*Note.* The top five and the least five countries reporting health events to WHO DON.

**Figure 2.**
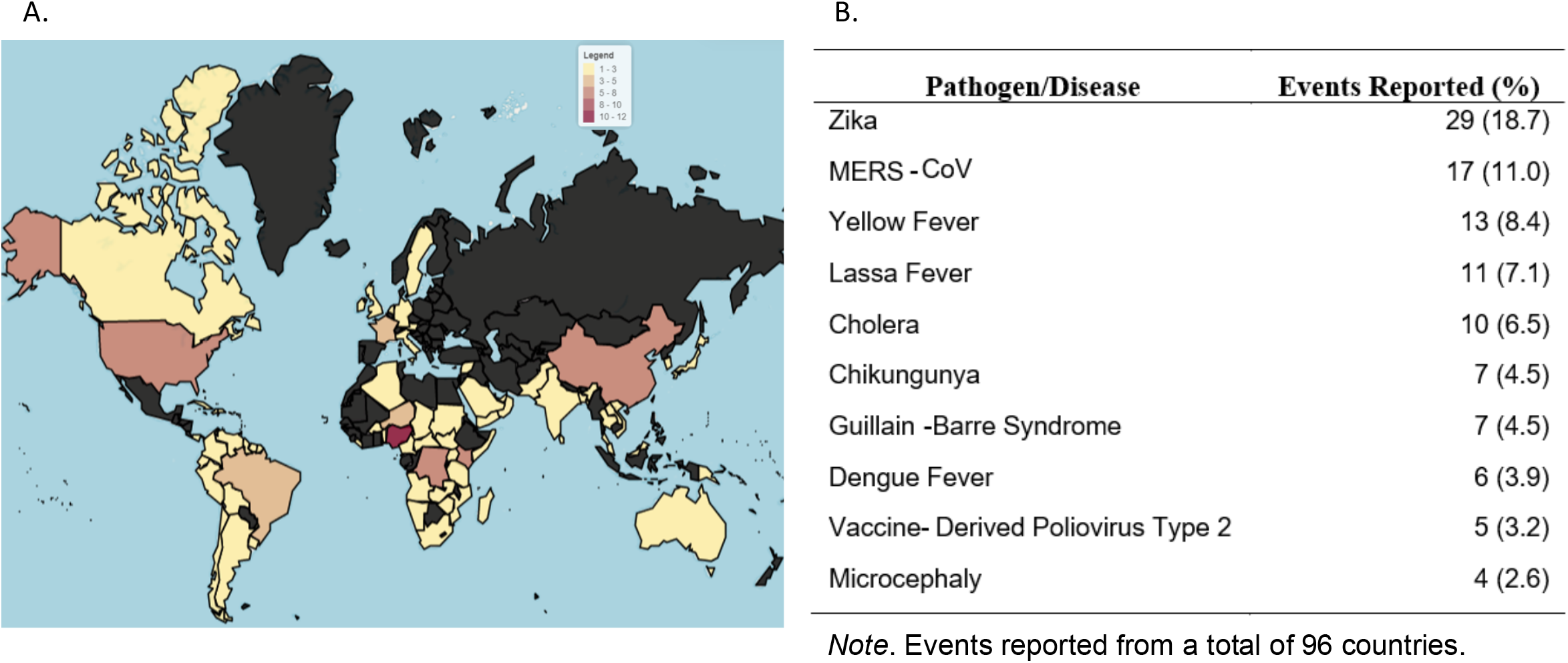
Major Health Event’s distribution by country and pathogens/disease reported on WHO Disease Outbreak News (2016-2018). **a**. Geographic heat map displaying the frequency of health events reported by country. **b**. Frequency of the main pathogens/diseases reported.

Extrinsic factors tested against health event frequency are described in Table 2. Total population and GDP per capital had the greatest magnitude of change. The least changing factor was female population with an overall mean change of -0.17 %. Bivariate Poisson models results showed that the extrinsic factors GDP per capita change, urban population change, and current average forest area were significantly associated with health event frequency (p=0.000, 0.004, and 0.000, respectively). Net migration change factor was the least related factor (p=0.838).

**Table 2.** Description of the extrinsic factors evaluated

| Extrinsic Factors | 2006-2008 | 2016-2018 | Mean Change |
| --- | --- | --- | --- |
|  | Mean (SD) | Mean (SD) |  |
| GDP per capita ( US 100 dollars) | 111.77 (167.82) | 120.73 (167.02) | 8.96* |
| Population ( million people) | 59.28 (191.57) | 66.43 (208.08) | 7.14* |
| Urban Population (%) | 54.66 (24.15) | 57.96 (23.84) | 3.30* |
| Female Population (%) | 49.43 (3.94) | 49.26 (4.52) | -0.17 |
| Population Density (people per sq. km of land area) | 186.32 (266.45) | 215.22 (324.09) | 28.90* |
| Net Migration | 6428.69 ( 993765.53) | -15462.46 (744692.31) | -21127.50 |
| Forest Area (million sq.km) | 32.32 (25.31) | 32.10 (25.47) | -0.39 |
| Trade (%) | 90.06 (78.08) | 76.38 (43.10) | -16.60* |
| Age 65 Older ( years) | 6.26 (4.59) | 7.42 (5.70) | 1.16* |

†Net migration data was available from 2007 and 2017. \*Paired T-test significant at $p < 0.05$ . N=96 countries from 2016-2018.

After testing all covariates (nine in total) against the dependent variable: health event frequency, multiple models were constructed. The final Poisson model includes three variables associated significantly to event frequency (population urban change, GDP change per capita, and the average forest area (2016-2018 current average)) and a fourth variable (percent of trade change), which is not statistically significant alone in the model but appears to interact with the other covariates (Table 3). The model suggests that an increase in the GDP change increases the likelihood of an event by 1% (p=0.00), increase in urban population change raises the likelihood by 7.2% (p=0.011), and an increase in average forest area (sq. meter) increases the likelihood by 27.9% (p=0.002). Model adequacy obtained from the goodness-of-fit Pearson chi-square was 0.850, suggesting a good fit of our data to a Poisson distribution in the regression. The likelihood ratio chi – square for model effects showed GDP change per capita, population urban change and average forest area have an discernible effect (p<0.05).

**Table 3.** Poisson regression model parameters estimates for health event frequency

| Explanatory Variable | Rate Ratio | 95% Confidence Interval for Rate Ratio |  | Significance Level (p- value) |
| --- | --- | --- | --- | --- |
| (Intercept) | 1.024 | 0.740 | 1.418 | 0.885 |
| GPD change per capita ( US 100 dollars) | 1.010 | 1.006 | 1.015 | <0.001* |
| Population urban change (%) | 1.072 | 1.016 | 1.132 | 0.011* |
| Average forest area sq. meter (million sq.km) | 1.279 | 1.095 | 1.494 | 0.002* |
| Trade percent change (%) | 1.000 | 0.997 | 1.003 | 0.932 |
| *Note.* Dependent Variable: Event Frequency. * p-value <0.05<br>N=96 countries from 2016-2018. |  |  |  |  |

Results related to model performance continued to show strong correlations with GDP change per capita, population urban change and average forest area in Monte Carlo simulations. A total of 71,075 cases were simulated using Monte Carlo method to meet the confidence interval of the mean of the target variables (health events), at the 95% confidence level. The tornado chart (Figure 3a) shows a strong Pearson correlation between health events and GDP per capita change (adjusted) of 0.85 and moderate correlations for the input variables population urban change and current average forest area (adjusted) with correlation coefficients of 0.54 and 0.40, respectively. The probability density chart (Figure 3b) displayed the distribution of the target variable (health events frequency) simulated by Monte Carlo Method. Results show a probability of 14% to have more than two health events and a probability of 87.9% to have at least one health event with our Poisson model indicators.

**Figure 3:**
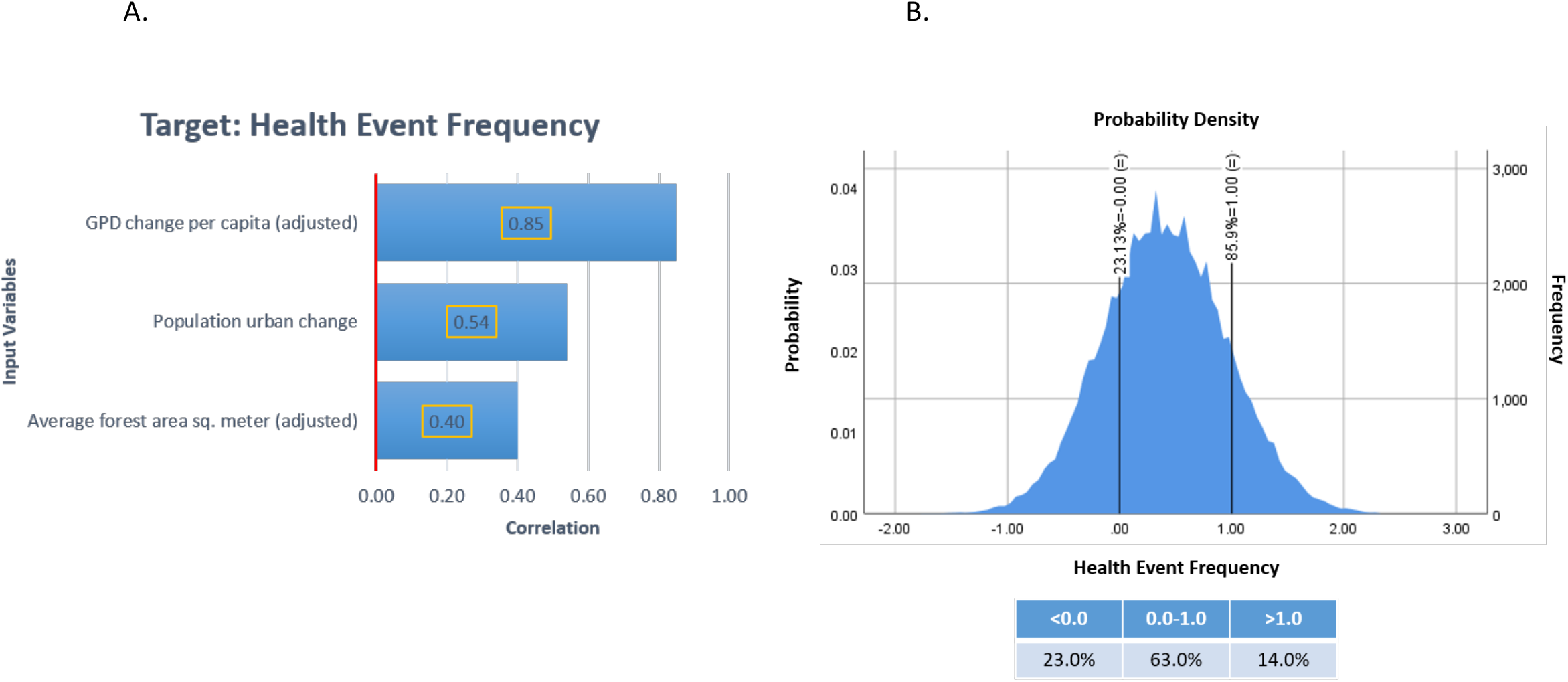
Monte Carlo simulation output using Poisson equation regression model. **a**. Correlation Tornado chart for input variables: GDP change per capita, population change and average forest area. **b**. Probability density chart of health event frequency; reference probability values were placed at 0.0 and 1.0 events.

A confounding analysis related to economic reporting behaviors was performed. It showed Venezuela, Nigeria, and the United States with the most GDP change; and, Haiti, Bahrain, and Pakistan with the least change. Among the low- and middle-income countries, global health security funds commitments were comparable for size with the exception of Venezuela; however, given the political distance between the major global health security donors and that country during the time period, this was not surprising. After evaluating comparisons among countries, an economic reporting bias was not found.

In light of the on-going novel coronavirus disease (COVID-19) public health emergency of international concern (PHEIC), we undertook an additional exploratory analysis of current COVID-19 cases reported in the countries included in our dataset. Two of our non-health indicators are also significantly correlated with COVID-19 reported cases. The correlation analysis showed GDP change per capita (p= 0.024) and average forest area (p<0.001) linkages to COVID-19 case counts. This is further supported with a linear regression model demonstrating that GDP change per capita, average forest area, and urban population change influence the outcome variable, in this case, COVID-19 cases reported (p<0.001). An R=0.5 suggests moderate correlation, and a 32% variance proportion in the dependent variable (COVID-19 reported cases, R2) can be predicted from these same three variable.

## Discussion

Research that pushes how surveillance and related activities happen is important.(6) Doing so can be challenging for many reasons. Medical intelligence and surveillance are increasingly multisectoral in nature resulting in many stakeholders which both compete and collaborate. Nonetheless, opportunities for enhancing public health practice exist in terms of increased scientific rigor, outcomes-focused research, and health informatics.(6) We sought a novel application of unconventional data for these purposes, selecting the WHO Disease Outbreak News because the events matter to communities, and the World Bank development factors as they reflect large aspects of community wellness. We applied them holistically in a way which mitigates their internal validity issues.

This regression model only includes those WHO countries which reported events during 2016-2018, resting on premises that past experiences are related to future experiences, and that these countries offer lessons for those which have not yet reported such events, even if they may have experienced analogous ones.

Our study evaluated a total of nine extrinsic factors; four of these were significantly related to frequency of a reported emerging infectious diseases event, with the average forest area sq. meter being the extrinsic factor with the highest effect, as shown in the Poisson model. Our simulation model showed that there is an 87% probability that a country will experience a health event during a three year observation period if it has undergone significant GDP, trade, and population changes in the previous 10 years while having a large forest area. Moving forward, intrinsic factors related to the health event must be incorporated, or a tiered approach adopted, in order to have a fuller picture of community and patient vulnerabilities for an event.

Previous studies have showed that extrinsic factors are associated with health; such as green space area, population density, wealth, education and others.(7-10) Our results showed an association between forest area and health events. More forest area available was associated with an increasing risk of a reported emerging infectious diseases event. Changes in land cover and land use, including forest area change (particularly deforestation and forest fragmentation), urbanization (which is included in our model as population urban change), and agricultural intensification are major factors contributing to the surge in infectious diseases.(11) A recent study sought to forecast the next forest-based emerging infectious disease and concluded that southern and eastern forests around Freetown in Sierra Leone, the forest region around Douala in Cameroon, or the southern forest region in Nigeria were potential upcoming originating centers of emerging infectious disease.(12) These regions have extensive forest areas, consistent with our model forecast, presumably from increased opportunities for zoonotic cross-over events. Exposure time is a dominant feature of risk.

Urban population change had a moderate association to health events in our model. In 2014, WHO asserted that urban areas held 54% of the total global population.(13) This ratio is expected to increase from 55% in 2018 (some 4.2 billion people) to 68% by 2050.(14) Our findings suggest that undergoing an urban population change increases risk of a health event. Whether this is better explained by forest incursion from urbanization; human consolidation and so increased human: human exposure time; aspects of domestic travel and draw from more wild fringe areas prone to sentinel zoonotic events; or, other root causes is less clear. Incorporation of more local market and migration factors could assist in making these distinctions. Regardless, with rapid global urbanization, understanding relationships between the changing urban environment and human health is vital. Urban environments play an indispensable role in influencing human health and wellbeing.(15)

Human population density has been recognized as a putative driver of emerging infectious diseases.(16) Human behavioral changes regarding movement and urbanization are thought to contribute to this.(17) Megacities may serve as incubators for new epidemics and zoonotic diseases of rapid spread.(18) Sporadic encounters between wildlife and humans in urban areas may become more frequent in peri-urban settings resulting in greater exposures to parasites, Dengue virus, cholera, tuberculosis, Lyme, and other threats.(16, 17, 19-23)

Our statistically strongest association rested with whether the gross domestic product of a country had undergone change in the intervening decade. Studies have shown associations between GDP and health outcomes, as well as increased health care expenditures and GDP growth.(24, 25) We specifically tested whether a changing GDP changed risk for a health event. This bore from our suspicion that whether increasing or decreasing, a changing GDP indicated a condition under which communities must adapt in ways that seek new markets and ways of instigating commerce, sporadic and sustained forest contact, and evolving interactions with urban areas. Indeed, our results demonstrate the novel risk indicator that actual increasing GDP change per capita increases the probability of an emerging infectious diseases event. This suggests that all countries are vulnerable to this effect regardless of baseline GDP.

There are new initiatives aimed at how risk models for infectious diseases are developed, such as the Epidemic Prediction Initiative from the Centers for Disease Control and Prevention.(26) While this initiative is meant to facilitate open forecasting projects towards public health decision making, they are targeted to explore specific pathogens, diseases, or vectors (e.g. Dengue, influenza, *Aedes aegypti*, and *Aedes albopictus*).(26) Our approach may be particularly useful for whole-of-society planning and operational health emergency risk management processes, such as called upon in the Joint External Evaluation monitoring progress in country attainment of IHR 2005 capacities, as well as other capability development and exercise initiatives.(27)

The main advantages of our approach are model simplicity; ready hypothesis generation from existing open source, long-lived data mechanisms; and, the ability to be adapted to specific epidemiological scenarios including contextual layering on current pathogen-specific prediction models. Employing non-health factors may improve a variety of models’ accuracy when predicting risks from pandemic threats. It offers readily observable triggers for public health strategic decision making and resource allocation; and, it may improve event-based surveillance. (28)

Our exploration into these effects’s impact on COVID-19 risk were limited. However, these results suggest that COVID-19 disease dynamics (e.g. outbreak magnitude, cases, death toll) when reported could be affected by the non-health indicators identified in our findings. The analysis may have heavily confounding by the pervasiveness of COVID-19 in the pandemic setting.

Our main limitations revolve around data collection in low resourced settings and reporting in politically associated systems. Variability existed in how data was reported in the WHO Disease Outbreak News. Demographical characteristics and epidemiological indicators were sometimes non-specific or missing. Under-reporting of cases occurred. For example, while Ebola virus disease, influenza, and Zika events were reported when the outbreak started in the country, subsequent reports of the outbreaks were omitted or contained limited information. Data extraction was made from WHO DON reports, therefore all the countries incorporated in analysis have at least one event reported. On one hand this introduces bias and limits the applicability of the findings to the non-reporting countries on the studied timeframe, on the other hand countries without a report at all were relatively few and the presence of at least one report indicates that the country will and has the mechanism to report. While our confounding analysis regarding country wealth, funding, and reporting behaviors yielded reassuring results, confounding may still be present.

## Conclusion

Our exploratory analysis demonstrates that non-health factors are associated with whether a country experiences a significant health event from an emerging or re-emerging infectious disease, based upon major health events (e.g. infectious diseases outbreaks) reported by WHO state members between 2016-2018. In addition to urban population, forest area, and trade, we identified a novel factor, GDP per capita change, which had the statistically strongest association in our model. Further exploration of dynamic non-health extrinsic factors as tools in prediction of such events anchored in community-centered outcomes is merited. Even now, the model may be useful in use case applications to contextualize whole-of-society threat planning processes and medical intelligence leading to public health decision-making for priority surveillance and other health and non-health investments.

## Data Availability

Database was developed using WHO Disease Outbreak News Reports from 2016-2018 (https://www.who.int/csr/don/en/) and economic variables were obtained from World Bank Open Data 2019 (https://data.worldbank.org/). Line listing is available to request.

## Acknowledgements

We are grateful to Dr. Richard Haas, economist and trade diplomat, for his insight on international economy and assistance regarding wealth indicators.

## Author contributions

DMB—project idea and design, data analysis, and content writing and editing. RLR—project design, data analysis, and content writing and editing. ECG—review of data analysis and content editing. CHO—review of data analysis.

## Disclaimer

The contents, views or opinions expressed in this publication are those of the authors and do not necessarily reflect official policy or position of Uniformed Services University of the Health Sciences, the Department of Defense (DoD), or Departments of the Army, Navy, or Air Force, or the State of Nebraska. Mention of trade names, commercial products, or organizations does not imply endorsement by the U.S. Government.

## Biographical Sketch

Clinical epidemiologist with experience in tropical infectious diseases such as Zika and Dengue.

## Current position

Clinical Epidemiologist Postdoctoral Fellow in the Preventive Medicine and Biostatics Department.

## Affiliations

Uniformed Services University, Bethesda; Henry M. Jackson Foundation, Bethesda

## Primary research area

Tropical and emerging diseases

## References

1. Rivers C, Chretien JP, Riley S, Pavlin JA, Woodward A, Brett-Major D, et al. Using “outbreak science” to strengthen the use of models during epidemics. Nat Commun. 2019;10(1):3102. Epub 2019/07/17. doi: 10.1038/s41467-019-11067-2. PubMed PMID:31308372; PubMed Central PMCID: PMCPMC6629683.

2. Organization. WH. International Health Regulations (2005). In: Organization WH, editor. Second ed. Switzerland 2008.

3. Scarpino SV, Petri G. On the predictability of infectious disease outbreaks. Nat Commun. 2019;10(1):898. doi: 10.1038/s41467-019-08616-0. PubMed PMID: 30796206; PubMed Central PMCID: PMCPMC6385200.

4. World Bank Group. World Bank Open Data 2019 [cited 2019]. Available from: https://data.worldbank.org/.

5. GIDA (Georgetown Infectious Disease Atlas). Recipients by country Committed funds. Georgetown University; 2019.

6. Henderson DA. The Development of Surveillance Systems. Am J Epidemiol. 2016;183(5):381–6. doi: 10.1093/aje/kwv229. PubMed PMID: 26928219.

7. Schiller JS, J. W. Lucas, J. A. Peregoy. Summary Health Statistics for U.S. Adults: National Health Interview Survey, 2011. In: Statistics VaH, editor.: Centers for Disease Control and Prevention 2012.

8. Pollack CE, Chideya S, Cubbin C, Williams B, Dekker M, Braveman P. Should Health Studies Measure Wealth?: A Systematic Review. American Journal of Preventive Medicine. 2007;33(3):250–64. doi: 10.1016/j.amepre.2007.04.033.

9. Maas J, Verheij RA, Groenewegen PP, de Vries S, Spreeuwenberg P. Green space, urbanity, and health: how strong is the relation? Journal of Epidemiology & Community Health. 2006 60:587–92.

10. Wolch JR, Byrne J, Newell JP. Urban green space, public health, and environmental justice: The challenge of making cities ‘just green enough’. Landscape and Urban Planning. 2014;125:234–44. doi: 10.1016/j.landurbplan.2014.01.017.

11. Wilcox BA, Ellis B. Forests and emerging infectious diseases of humans,. Unasylva. 2006;57(224):11–8.

12. Shah V, Shah A, Joshi V. Predicting the origins of next forest-based emerging infectious disease. Environ Monit Assess. 2018;190(6):337. Epub 2018/05/11. doi: 10.1007/s10661-018-6711-6. PubMed PMID: 29744690.

13. World Health Organization. Global Health Observatory (GHO) data 2019 [cited 2019]. Available from: https://www.who.int/gho/urban_health/situation_trends/urban_population_growth_text/en/.

14. United Nations DoEaSA, Population Division. World Urbanization Prospects 2018: Highlights New York: UNITED NATIONS; 2019 [cited 2019]. Available from: https://population.un.org/wup/Publications/Files/WUP2018-Highlights.pdf.

15. Bai X, Nath I, Capon A, Hasan N, Jaron D. Health and wellbeing in the changing urban environment: complex challenges, scientific responses, and the way forward. Current Opinion in Environmental Sustainability. 2012;4(4):465–72. doi: 10.1016/j.cosust.2012.09.009.

16. Daszak P, Cunningham AA, Hyatt AD. Anthropogenic environmental change and the emergence of infectious diseases in wildlife. Acta Tropica. 2001;78(2):103–16. doi: 10.1016/s0001-706x(00)00179-0.

17. Morens DM, Folkers GK, Fauci AS. The challenge of emerging and re-emerging infectious diseases. Nature. 2004;430(6996):242–9. doi: 10.1038/nature02759.

18. Neiderud CJ. How urbanization affects the epidemiology of emerging infectious diseases. Infect Ecol Epidemiol. 2015;5:27060. Epub 2015/06/27. doi: 10.3402/iee.v5.27060. PubMed PMID: 26112265; PubMed Central PMCID: PMCPMC4481042.

19. Polley L. Navigating parasite webs and parasite flow: emerging and re-emerging parasitic zoonoses of wildlife origin. Int J Parasitol. 2005;35(11-12):1279-94. Epub 2005/09/20. doi: 10.1016/j.ijpara.2005.07.003. PubMed PMID: 16168994.

20. Teixeira MdG, Barreto ML, Costa MdCN, Ferreira LDA, Vasconcelos PFC, Cairncross S. Dynamics of dengue virus circulation: a silent epidemic in a complex urban area. Tropical Medicine and International Health. 2002;7(9):757–62. doi: 10.1046/j.1365-3156.2002.00930.x.

21. Gubler DJ. Dengue, Urbanization and Globalization: The Unholy Trinity of the 21(st) Century. Trop Med Health. 2011;39(4 Suppl):3-11. Epub 2012/04/14. doi: 10.2149/tmh.2011-S05. PubMed PMID: 22500131; PubMed Central PMCID: PMCPMC3317603.

22. Dunkle SE, Mba-Jonas A, Loharikar A, Fouche B, Peck M, Ayers T, et al. Epidemic cholera in a crowded urban environment, Port-au-Prince, Haiti. Emerg Infect Dis. 2011;17(11):2143-6. Epub 2011/11/22. doi: 10.3201/eid1711.110772. PubMed PMID: 22099120; PubMed Central PMCID: PMCPMC3310575.

23. Hayward AC, Darton T, Van-Tam JN, Watson JM, Coker R, Schwoebel V. Epidemiology and control of tuberculosis in Western European cities. Int J Tuberc Lung Dis. 2003;7(8):751-7. Epub 2003/08/19. PubMed PMID: 12921151.

24. Fuchs VR. The gross domestic product and health care spending. N Engl J Med. 2013;369(2):107-9. Epub 2013/05/24. doi: 10.1056/NEJMp1305298. PubMed PMID: 23697470.

25. Bradley E, Canavan M, Rogan E, Talbert-Slagle K, Ndumele C, Taylor L, et al. Variation In Health Outcomes: The Role Of Spending On Social Services, Public Health, And Health Care, 2000–09. Health Affairs. 2016;35(5):760–8. doi: https://doi.org/10.1377/hlthaff.2015.0814.

26. Centers for Disease Control and Prevention. Epidemic Prediction Initiative (BETA). 2019.

27. Alliance for Country Assessments for Global Health Security and IHR Implementation. Joint External Evaluation (JEE). Geneva, Switzerland 2019.

28. World Health Organization. Early detection, assessment and response to acute public health events: Implementation of Early Warning and Response with a focus on Event-Based Surveillance. Geneva: World Health Organization; 2014. p. 1–59.

